# Community knowledge, attitudes and practices related to Taenia solium taeniosis and cysticercosis in Zambia

**DOI:** 10.1101/2023.05.14.23289972

**Authors:** Gideon Zulu, Kabemba E. Mwape, Tamara M. Welte, Martin Simunza, Alex Hachangu, Wilbroad Mutale, Mwelwa Chembensofu, Chummy Sikasunge, Isaac K. Phiri, Andrea S. Winkler

**Author notes:** Corresponding author (GZ). ^#^ These authors contributed equally to this work (Isaac K. Phiri, Andrea S. Winkler).

## Abstract

**Background:** Several studies on *Taenia solium* taeniosis / cysticercosis (TSTC) have been conducted in Zambia. However, none has assessed community knowledge, attitudes and practices related to TSTC and epilepsy.

**Methods:** A community-based cross-sectional study was conducted between November and December 2022. The design consisted of a questionnaire-based survey conducted in each of the 25 purposely selected villages in Chiparamba Rural Health Centre (RHC) catchment area in Chipata district of the Eastern Province.

**Results:** A total of 588 participants comprising 259 (44%) males and 329 (56%) females with median age of 42 years (range 17 to 92 years) were interviewed. Awareness of the signs and symptoms of taeniosis and human cysticercosis (HCC), including transmission and prevention measures was very low. Whilst the majority had heard about epilepsy, they were not able to link HCC to epilepsy. Most participants were aware of cysticerci in pigs (PCC) including its predilection sites but were not aware of mode of transmission and prevention measures. The pork meat inspection by trained professionals was also not a common practice in the area. Risk perception of *T. solium* infections was thus very low. Overall knowledge, attitude and practice scores related to *T. solium* infections and to epilepsy were very low with median scores of 0.38 (IQR 0.25 - 0.54) for knowledge, 0.25 (0.25 - 0.50) for attitudes, and 0.31 (0.25 - 0.44) for practices. Males had better knowledge on TSTC (median = 0.42, p = 0.017, r = 0.098) and better practice scores (median = 0.38, p = < 0.001, r = 0.154) compared to females though the effect size was small. With regards to sanitation and hygiene washing with soap and water was reported by many but only few had a hand washing facility near their latrines.

**Conclusion:** The study shows overall poor knowledge, attitudes and practices related to TSTC among the community of Chiparamba RHC in Chipata district of the Eastern Province of Zambia. This poses a serious challenge for control and elimination of *T. solium* infections and thus efforts to improve knowledge, attitudes and practices should be made using a One Health approach for the control and elimination of TSTC. Educational programs about TSTC transmission, signs and symptoms, prevention, management and control need to be scaled up in the study area and Zambia as a whole.

## Author summary

*Taenia solium* taeniosis / cysticercosis (TSTC) is endemic in Zambia. We aimed at assessing knowledge, attitudes and practices (KAP) related to TSTC and epilepsy in a community setting in Chiparamba area of Chipata district of the Eastern Province of Zambia. Understanding KAP is important in designing and implementing public health interventions for control and eradication of TSTC in a community. We found generally poor KAP related to TSTC among the community in our study area. This lack of knowledge about TSTC infections can lead to behaviours that facilitate the transmission and maintenance of *T. solium* infections and could hinder efforts to control the most preventable cause of epilepsy in the sub-Saharan African region. More efforts to improve KAP using a One Health approach for the control and elimination of TSTC are therefore required in Zambia.

## Introduction

*Taenia solium* is a neglected zoonotic tapeworm causing taeniosis in humans and cysticercosis in pigs and humans. Taeniosis, which is the presence of an adult tapeworm within the human intestine, occurs when one ingests undercooked pork infected with *T. solium* cysticerci. The adult worm develops and sheds mature proglottids containing thousands of eggs through the faeces of a *T. solium* carrying individual (definitive host). Pigs through their coprophagic nature get infected with *T. solium* by consuming the infected faeces from a *T. solium* carrying individual. Once the eggs are ingested by pigs, they hatch into oncospheres in the intestines and migrate to the muscle and other tissues within the pig causing porcine cysticercosis (PCC) (intermediate host) and thus completing the life cycle. Humans can also develop cysticercosis (HCC) if they ingest *T. solium* eggs through autoinfection (for a tapeworm carrier) or through contaminated water or food (faecal-oral). The larvae migrate through the circulatory system to various tissues causing cysticercosis, but if they lodge in the central nervous system (CNS), then neurocysticercosis (NCC) may occur (1). Taeniosis is mostly asymptomatic but signs / symptoms vary from abdominal discomfort, vomiting and the passage of visible tapeworm proglottids in stool. Cysticercosis is difficult to diagnose but may show subcutaneous nodules for those with cysticerci in the subcutaneous tissues (2,3). NCC may also remain asymptomatic but some present with severe progressive headache, and / or epileptic seizures, sometimes also with focal neurological signs / symptoms (4). *T. solium* is among the list of foodborne zoonotic parasites causing considerable morbidity and mortality and is the leading cause of epilepsy and other neurological sequelae in *T. solium* endemic populations across sub-Saharan Africa (5–7).

In many low-income and middle-income countries (LMICs), *T. solium* cysticercosis is a neglected zoonotic disease in humans and pigs and is underreported (8). *T. solium* taeniosis / cysticercosis (TSTC) has significant public health and economic impacts, although the exact impact is difficult to determine due to lack of meat inspection to detect the cysts, backyard slaughter of pigs, lack of affordable diagnostic tools in humans and stigma related to NCC in endemic areas (9–11). *T. solium* cysticercosis prevention and control measures require a good understanding about the transmission dynamics and the factors that perpetuate its presence in the communities.

In Zambia, a number of studies on *T. solium* infections have been conducted (12,13). These include studies assessing community perceptions on the use of latrines (14,15), pig management practices (16) and an assessment of acceptability of control measures for *T. solium* infections (16). However, no study has assessed community understanding of the relationship between taeniosis, cysticercosis (in pigs and humans) and NCC as well as NCC and epilepsy. This study aimed at assessing the knowledge, attitudes and practices (KAP) related to TSTC and NCC in Chipata district in Eastern Province of Zambia.

The study was done according to the strobe checklist (S1 Table).

## Materials and Methods

### Ethics

Ethical clearance was obtained from ERES CONVERGE Institutional Review Board (IRB), reference number 2018-March-002. Approval was also obtained from the Zambia National Health Research Authority (ZNHRA). All participants were informed about all aspects of the study before inclusion, and all signed an informed consent.

The reporting of this study followed the STROBE checklist (S1 Table).

### Study area

The study was conducted in Chipata district in the Eastern Province of Zambia within the Chiparamba Rural Health Centre (RHC) catchment area with 25 selected villages. The RHC has a catchment population of 17,881 (clinic head count). The main economic activity for the people in this area is subsistence farming especially growing maize and groundnuts for home consumption. The community keeps pigs, goats and chickens as their main livestock. The main ethno-linguistic group of the area are the Chewas and Ngonis. The Chewas being of a matrilineal descent while the Ngonis are of a patrilineal descent. The area was selected because it is well known to the researchers for practicing free-range pig keeping, low levels of sanitation and a high number of epilepsy patients reporting to health facilities within the district, as well as its proximity to a neighbouring Katete district with high *T. solium* cysticercosis prevalence in pigs and humans (18,19)

### Study design

This study forms part of a larger study (CYSTINET-Africa) related to the occurrence and detection of human *T. solium* infections in the Eastern Province of Zambia. A community-based cross-sectional study was conducted between November and December 2022. The design consisted of a questionnaire-based survey conducted in each of the 25 purposely selected villages in Chiparamba RHC catchment area. A list of all households was available from an ongoing study about *T. solium* in the district. The households were randomly selected from each village. In each of the selected households, the interviews were conducted with the head of the household and in case the head was absent, another adult household member was interviewed.

To ensure that correct information was obtained when referring to a “tapeworm” and “cysticerci” and to reduce the possibility of confusing these with other parasites, a colour picture of an adult tapeworm and another of pork infested with cysticerci were shown to the respondents during the interview (S1 File: S1 Fig).

### Sample size determination

The sample size calculation was performed using the formula for qualitative variables with an infinite population (20). Admitting no previous knowledge about TSTC in the study population (p = 0.5), with a significance level of 95% (Z α/2 = 1.96) and a margin error of 5%, the minimum sample size calculated was 384. A total number of 588 households, originating from 25 villages of Chiparamba RHC catchment area, were included to cater for non-response and 50% representation from each of the randomly selected villages.

### Questionnaire survey

We adopted a questionnaire which was validated and used in Tanzania for a similar study (21). The questionnaire underwent a validation process by a team of experts (ASW, IKP, KEM, TW and WM) regarding objectivity, clarity and relevance. Before data collection, content validity of the questionnaire was discussed among these experts to assess conformity of the questionnaire to the study objective and to obtain consensus. Face validity was assessed by piloting the questionnaire on ten respondents from a non-participating village within the RHC catchment area to assess understanding of the questions and whether they were easy to follow. The questions were phrased according to the local circumstances and face-to-face interviews were used to administer the questionnaire to each of the selected household members by a team comprising six investigators conversant in the local language (Nsenga) common to both the Chewa and the Ngoni people. The questionnaire contained 87 questions altogether (S2 File) divided into eight sections. Section one comprised six questions on general information (date of interview, district name, name of village and household number, names of household head and interviewer). Section two of the questionnaire had seven questions about personal information of the respondent. Section three comprised thirteen questions about drinking water and sanitation. Section four comprised nine questions regarding human tapeworm infection. Section five comprised twelve questions regarding HCC infection which also included questions regarding NCC. Section six comprised ten questions regarding information on pork consumption and management. Section seven comprised thirteen questions regarding PCC and section eight comprised seventeen questions regarding knowledge, attitudes and perceptions related to epilepsy. The presence and quality of toilets, presence of hand washing facilities inside or outside the latrines, presence of a dish rack and pig management systems (confinement, tethering, or free-range) were assessed through direct observations. All interviews were conducted in Nsenga the local language in the Chiparamba area of Chipata district.

### Data collection

Data was collected using KoboCollect, an application for data collection installed on tablets (22). The investigators were trained on the use of the KoboCollect tool before commencement of the study and the feasibility and correctness of the items in the electronic questionnaire was also pre-tested.

### Study variables

The terms knowledge, attitudes and practices were defined according to Badran (23). Within the context of our study knowledge refers to the capacity to acquire and retain information about *T. solium* infections (*T. solium* taeniosis, HCC, NCC and PCC) and epilepsy. Attitude refers to inclinations to react in a certain way towards *T. solium* infections including epilepsy and by practice we mean the application of rules and knowledge that leads to actions regarding *T. solium* infections and epilepsy. Participants that had heard about *T. solium* infections and epilepsy were described as “being aware” of these conditions. Within the aspects of KAP, a minimum score of 0 points and a maximum score of 1 point was allocated to each question in the questionnaire. In questions with more than one correct alternative, scores were considered proportional to the number of correct answers. Final KAP scores were defined as the arithmetic mean of the scores within each section of the KAP, i.e. knowledge, attitudes and practices separately per individual. The scores were thus represented by quantitative variables (dependent), described within the closed interval between 0 and 1, where the higher the score value, the higher the evaluation of the respondent’s KAP.

### Statistical analysis

Data were stored in a Microsoft Excel 2016 database (Microsoft Corporation; Redmond, WA, EUA) where scores of each component were calculated. Later, statistical analyses were performed using IBM SPSS Statistics (Version 23). Absolute and relative (%) frequencies were calculated for descriptive analyses of the independent variables. For the dependent variables in the descriptive analysis, median, interquartile range, and minimum and maximum values were calculated. The non-parametric Mann-Whitney-U and Kruskal-Wallis tests were performed to determine associations of the scores with independent variables (24). The effect size (r-statistic), was calculated and values between 0.1 and 0.3 were classified as a small effect, between 0.3 and 0.5 were classified as a moderate effect and greater than or equal to 0.5 were classified as a large effect (25). To evaluate the relationship between the scores, Spearman correlation was applied, and a Kruskal-Wallis test was used to compare the scores (24). Associations between categorical variables were assessed using the Chi-square test and a *p*-value less or equal to 0.05 was considered to be significant.

## Results

In total, 588 interviews from distinct households were conducted. The study included 259 (44%) male and 329 (56%) female participants. The age ranged from 17 to 92 years (median 42 years) with the majority being older than 30 years. Fig 1 shows the age distribution of the respondents indicating that a good number (35%) was above fifty (50) years of age.

**Fig 1.**
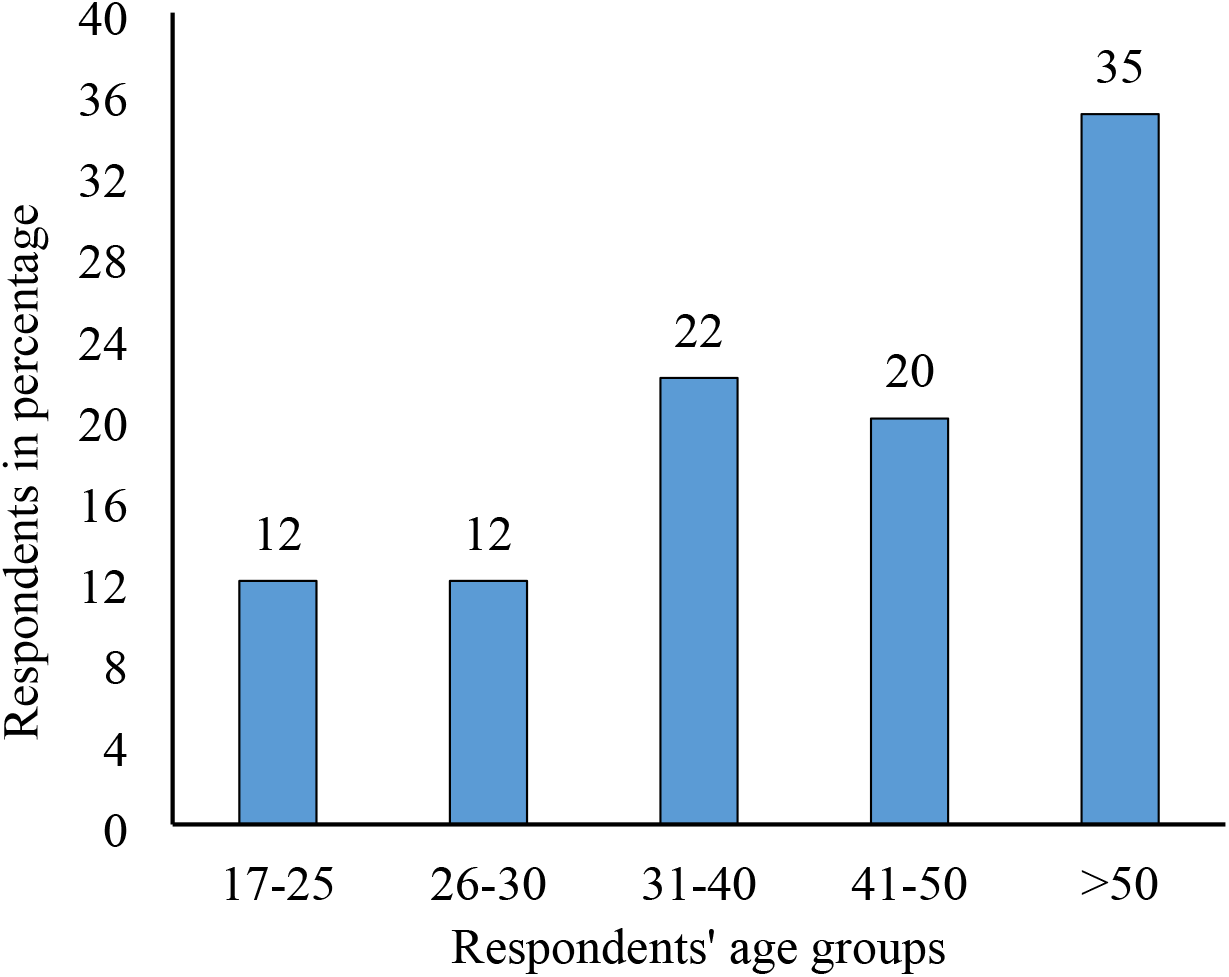
Age group classification of respondents (N = 588) of Chiparamba in Chipata district.

Most of the participants were of the Christian faith, predominantly Catholic and Protestant with only 28 (5%) comprising participants of the Muslim faith, as shown in Table 1. Farming was the main occupation (557, 95%) as well as the main source of income for most households (547, 93%). On education, the majority of respondents (364, 62%) only went up to primary level with 132 (22%) having some secondary education (Table 1).

**Table 1.** Social demographic characteristics of participants (N = 588) of Chiparamba in Chipata district.

| Variable |  | Number (n) | Percent (%) |
| --- | --- | --- | --- |
| Sex | Male | 259 | 44 |
|  | Female | 329 | 56 |
| Age (years) | 17-25 | 69 | 12 |
|  | 26-30 | 69 | 12 |
|  | 31-40 | 131 | 22 |
|  | 41-50 | 115 | 20 |
|  | >50 | 204 | 35 |
| Religion | Catholic | 178 | 30 |
|  | Protestant | 337 | 57 |
|  | Adventist | 4 | 1 |
|  | Muslim | 28 | 5 |
|  | Other | 41 | 7 |
| Main occupation | Farmer | 557 | 95 |
|  | None | 6 | 1 |
|  | Other | 25 | 4 |
| Level of education | None | 91 | 15 |
|  | Primary school | 364 | 62 |
|  | Secondary school | 132 | 22 |
|  | Tertiary | 1 | 0 |
| Main source of income | Farming | 547 | 93 |
|  | Business | 28 | 5 |
|  | Formal employment | 1 | 0 |
|  | Other | 12 | 2 |
| Main occupation | Farmer | 557 | 95 |
|  | None | 6 | 1 |
|  | Other | 25 | 4 |

Sixty percent (351) of participants interviewed had heard about pork tapeworm infection in humans (*T. solium* taeniosis) and about half of these (163, 46%) first learned about it through community sensitization conducted before the start of the study, while 30% (107) heard about it from hospital staff (S1 File: S1 Table). Table 2 describes knowledge among participants that had heard (were aware) about *T. solium* taeniosis, HCC / NCC, PCC and epilepsy. Only 40% (141) of those who had heard about tapeworm infection were knowledgeable about the signs and symptoms of taeniosis. Knowledge about tapeworm transmission and prevention was equally low with only 16% (57) of participants being aware of infection through eating infected pork and 39% (138) giving correct responses about prevention. The majority of the participants (98, 28%) wrongly thought tapeworm was acquired from eating contaminated food (S1 File: S1 Table). Fifty-seven percent (334) of the respondents had heard about HCC but only 1% (4) of these were aware of the transmission through consumption of *T. solium* eggs. Similarly, only 6% (21) were aware of prevention of HCC. Most participants (215, 64%) were aware of the location of HCC lesions but knowledge regarding HCC and NCC signs / symptoms was low with only 28% (94) and 39% (131) of participants giving correct responses for HCC and NCC signs / symptoms respectively (Table 2, S1 File: S2 Table).

**Table 2.** Participant’s knowledge related to *T. solium* taeniosis / cysticercosis, porcine cysticercosis and epilepsy (n = 588).

| **Description** | **Number (n)** | **Percent (%)** |
| --- | --- | --- |
| ***Awareness of pork tapeworm in humans\**** | **351** | **60** |
| Signs of infection | 141 | 40 |
| Transmission of tapeworm | 57 | 16 |
| Tapeworm prevention | 138 | 39 |
| ***Awareness of human cysticercosis \**** | **334** | **57** |
| Cysticercosis transmission | 4 | 1 |
| Locations for cysticercosis lesions | 215 | 64 |
| Symptoms of cysticercosis | 94 | 28 |
| Signs of neurocysticercosis | 131 | 39 |
| Prevention of cysticercosis | 21 | 6 |
| ***Awareness of porcine cysticercosis\**** | **411** | **70** |
| Location of cysts in pigs | 376 | 91 |
| Transmission of porcine cysticercosis | 64 | 16 |
| Prevention of porcine cysticercosis | 179 | 44 |
| ***Awareness of epilepsy in humans*** | **566** | **96** |
| Linkage of epilepsy to human cysticercosis | 45 | 8 |

\*Responses within the groups are not mutually exclusive

Most of the participants (411, 70%) knew about cysticerci in pigs (PCC) with 91% (376) of them correctly identifying the location of cysts in muscles and under the tongue. However, knowledge regarding transmission of PCC was very low with only 16% (64) of those aware of PCC being knowledgeable on how pigs acquire cysticerci. Thirty-four percent (59) of those that said they knew how pigs acquired cysticerci believed that PCC was due to feeding of pigs with “gaga” the remains of maize bran that is used to make local beer (S1 File: S3 Table). Only few participants knew the transmission of PCC, consequently knowledge on prevention of PCC was very low with only 44% (179) giving the correct responses (Table 2).

The majority of the participants (566, 96%) had heard about epilepsy (Table 2) with 58% (328) knowing someone in the village who had epilepsy. However, very few (45, 8%) knew the association between epilepsy and HCC / NCC (S1 File: S4 Table).

Risk perception of *T. solium* tapeworm infection and cysticercosis was very low with only 39% (136) and 35% (116) believing that they were at risk of taeniosis and cysticercosis respectively (Table 3, S1 File: S5 Table). Regarding attitudes towards *T. solium* taeniosis prevention, most participants regarded pork infected with cysticerci as bad meat with 94% (385) reporting that they would not eat infected pork, 87% saying they would not sell the pork and 91% saying they would discard the infected pork (Table 3, S1 File: S5 Table). On assessment of attitudes towards epilepsy, the majority of participants (510, 90%) considered epilepsy a very serious disease and correctly indicated that people with epilepsy (PWE) should go to hospital for medical help. Similarly, most participants had correct attitude regarding where to seek help if one had tapeworm infection or HCC (Table 3).

**Table 3.** Attitudes and practices regarding *T. solium* taeniosis / cysticercosis, porcine cysticercosis and epilepsy.

| **Factor** | **Number (n)** | **Percent (%)** |
| --- | --- | --- |
| ***Risk perception of human tapeworm infections (n = 351) \**** |  |  |
| Believe they are at risk of infection with tapeworm | 136 | 39 |
| Would go to hospital if had tapeworm infection | 348 | 99 |
| ***Risk perception of human cysticercosis infections (n = 334) \**** |  |  |
| Believe they are at risk of cysticerci infection | 116 | 35 |
| Would go to hospital if had human cysticercosis | 314 | 94 |
| ***Attitudes towards infected pork (n = 411) \**** |  |  |
| Would sell pork with cysts | 52 | 13 |
| Would eat pork with cysts | 26 | 6 |
| Would discard pork with cysts | 374 | 91 |
| ***Attitudes regarding human epilepsy (n = 566) \**** |  |  |
| Considers epilepsy to be a serious disease | 510 | 90 |
| Would go to hospital if one has epilepsy | 560 | 99 |
| ***Pig rearing practices (n = 588)*** |  |  |
| Keeps pigs | 98 | 17 |
| *Free-range pig rearing practice* | 54 | 55 |
| *Permanently housed pigs* | 27 | 28 |
| *Pigs housed at night only* | 17 | 17 |
| ***Pig slaughtering practices (n = 588)*** |  |  |
| Slaughtered pig at home | 108 | 18 |
| *Meat inspected after slaughter* | 11 | 10 |
| ***Pork consumption practices (n = 588)*** |  |  |
| Eats pork meat | 420 | 71 |
| *Cook by boiling the pork meat* | 400 | 95 |
| ***Sanitation and hygiene practices (n = 588)\**** |  |  |
| Treat drinking water | 46 | 8 |
| Always wash hands after using the toilet | 419 | 71 |
| Wash hands using water with soap or ash | 305 | 52 |

\*Responses within factors are not mutually exclusive

Regarding practices and with respect to pig keeping, only 17% (98) of the participants kept pigs and 55% (54) of these raised their pigs through free-range method, 28% (27) permanently housed their pigs and 17% (17) housed them at night (Table 3). Of all participants interviewed, only 18 % (108) had slaughtered a pig at home and among these, only 10% (11) had their meat inspected. The majority (65, 67%) reported that there was no need for inspection of the meat while 22% (21) said there were no inspectors to conduct the inspection of the slaughtered pigs (S1 File: S6 Table). Seventy-one percent (420) of the participants consumed pork meat and 95% (400) of these preferred their meat prepared by boiling. Eighty-eight percent (517) of the participants had seen cysticerci in pork meat before and 37% (191) of them reported that the contaminated meat which they saw was sold, 20% (101) reported that it was eaten and 39% (202) reported that the meat was returned to the owner (S1 File: S6 Table).

Regarding sanitation and hygiene, the majority of the participants (550, 94%) got their drinking water from boreholes and most of them (542, 92%) did not treat their drinking water. Whilst most participants (419, 71%) reported washing their hands after using the toilet, only 52% (305) of these used water with soap or ash (Table 3, S1 File: S7 Table).

### Analysis of KAP scores

Analysing the KAP scores related to TSTC and epilepsy, it was observed that overall, all the scores were low. The knowledge variable obtained the highest score, whereas attitude had the lowest. When comparing the knowledge and practice scores, the two variables had a significant difference (p < 0.001) between them, with good knowledge scores being higher than the corresponding practice scores (Table 4).

**Table 4.** Scores for knowledge, attitudes and practices regarding *T. solium* taeniosis and cysticercosis of participants from Chiparamba in Chipata district.

|  | Spearman's rho |  |  | KAP scores |  |  |  |
| --- | --- | --- | --- | --- | --- | --- | --- |
|  | K | A | P | MD | IQR | Min | Max |
| **Knowledge** | 1.000 | 0.492** | 0.223** | 0.38 | 0.25 - 0.54 | 0.00 | 0.83 |
| **Attitude** | 0.492** | 1.000 | 0.243** | 0.25 | 0.25 - 0.50 | 0.00 | 1.00 |
| **Practice** | 0.223** | 0.243** | 1.000 | 0.31 | 0.25 - 0.44 | 0.00 | 0.69 |

K, Knowledge; A, Attitudes; P, Practices; MD, median score; IQR, Interquartile range; Min, Minimum score, Max, Maximum score
\*\* Correlation is significant at the 0.01 level (2-tailed).

The overall descriptive analysis of knowledge, attitude and practice scores showed that among the three components, the knowledge score (MD = 0.38) was higher than the attitude (MD = 0.25) and practice scores (MD = 0.31). Both comparisons were significantly different (*p* < 0.001). There was also a significant difference with a lower score for attitude when compared to the other variables (Table 4). The Spearman correlation between knowledge and attitude scores (r = 0.492; *p* < 0.001) was positive and significant, indicating that the higher the knowledge score, the higher the attitude score. The correlation coefficient between knowledge and practice scores (r = 0.223; *p* < 0.001) was also positive and significant, showing that the higher the knowledge score, the better the practice. Regarding the correlation between attitude and practice scores (r = 0.243; *p* < 0.001), it was positive and significant, indicating that the higher the attitude score, the better the practice score (Table 4).

Considering the association of KAP scores with the socio-demographic factors it was found that regarding the knowledge component, there was no statistically significant difference in knowledge on TSTC across the various age groups (*p* = 0.977). Neither was there any statistically significant difference in knowledge by religion (*p* = 0.059) or level of education (*p* = 0.160). However, it was observed that males had better knowledge on TSTC than females, though the effect size for this difference was small (MD = 0.42, *p* = 0.017, r = 0.098) (Table 5, S1 File: S8 Table). Regarding the attitude’s component, there was no statistically significant difference in attitudes towards TSTC within the variables age group (*p* = 0.698), sex (*p* = 0.112), religion (*p* = 0.177), and level of education (*p* = 0.88). Within the practice component, males had better practice scores than females, though the effect size was also small (MD = 0.38, *p* = < 0.001, r = 0.154). There was no statistically significant difference in practices between the various age categories (*p* = 0.267) though there was a tendency of better practices among the age group 17 - 24 years (MD = 0.38). Similarly, no statistically significant difference was observed in practices based on religion (*p* = 0.774) or level of education (*p* = 0.326) however, Catholics had better practices (MD = 0.38) compared to the other religions (Table 5).

**Table 5.** Association of KAP scores with socio-demographic factors.

|  |  | Knowledge |  |  |  | Attitude |  |  |  | Practice |  |  |  |
| --- | --- | --- | --- | --- | --- | --- | --- | --- | --- | --- | --- | --- | --- |
| Characteristic |  | MD | IQR | p-value | ES | MD | IQR | p-value | ES | MD | IQR | p-value | ES |
| Age (years) | 17-25 | 0.38 | 0.79 | 0.977* |  | 0.25 | 1.00 | 0.698* |  | 0.38 | 0.63 | 0.267* |  |
|  | 26-30 | 0.33 | 0.71 |  |  | 0.25 | 0.75 |  |  | 0.31 | 0.63 |  |  |
|  | 31-40 | 0.38 | 0.75 |  |  | 0.25 | 1.00 |  |  | 0.31 | 0.57 |  |  |
|  | 41-50 | 0.38 | 0.83 |  |  | 0.25 | 1.00 |  |  | 0.31 | 0.56 |  |  |
|  | >50 | 0.38 | 0.83 |  |  | 0.25 | 0.75 |  |  | 0.31 | 0.69 |  |  |
| Sex | Male | 0.42 | 0.83 | 0.017<sup>#</sup> | 0.098 | 0.25 | 1.00 | 0.112<sup>#</sup> |  | 0.38 | 0.69 | < 0.001<sup>#</sup> | 0.154 |
|  | Female | 0.33 | 0.83 |  |  | 0.25 | 1.00 |  |  | 0.31 | 0.69 |  |  |
| Religion | Catholic | 0.38 | 0.75 | 0.059* |  | 0.25 | 1.00 | 0.177* |  | 0.38 | 0.63 | 0.774* |  |
|  | Protestant | 0.38 | 0.83 |  |  | 0.25 | 1.00 |  |  | 0.31 | 0.69 |  |  |
|  | Adventist | 0.30 | 0.33 |  |  | 0.25 | 0.25 |  |  | 0.31 | 0.31 |  |  |
|  | Muslim | 0.29 | 0.75 |  |  | 0.25 | 0.75 |  |  | 0.31 | 0.50 |  |  |
|  | Other | 0.29 | 0.71 |  |  | 0.25 | 1.00 |  |  | 0.31 | 0.63 |  |  |
| Education | None | 0.38 | 0.71 | 0.160* |  | 0.25 | 0.75 | 0.880* |  | 0.31 | 0.63 | 0.326* |  |
|  | Primary | 0.38 | 0.83 |  |  | 0.25 | 1.00 |  |  | 0.38 | 0.69 |  |  |
|  | Secondary | 0.33 | 0.75 |  |  | 0.25 | 1.00 |  |  | 0.31 | 0.63 |  |  |
|  | Tertiary | 0.71 | 0.00 |  |  | 0.25 | 0.00 |  |  | 0.38 | 0.00 |  |  |

ES, Effect size (0.1 = Small ES, 0.3 = Medium ES, 0.5 = Large ES); MD, Median score; IQR, Interquartile range.
\**p*-values based on Kruskal-Wallis test; <sup>#</sup>*p*-values based on Mann-Whitney-U test .

## Discussion

This study conducted in the Eastern Province of Zambia, an area endemic for *T. solium* infections, showed that analyses of KAP was important to determine the vulnerabilities that surround knowledge, attitudes and practices of the population regarding this disease of public health importance.

Our findings indicate that the majority of the respondents do not have adequate knowledge about *T. solium* presence in their communities, signs / symptoms of TSTC, infection transmission, prevention and control. With the increase in pig farming in the region this lack of knowledge could lead to further increase of TSTC infections as the community is already exposed. The low knowledge levels about tapeworm infection amongst the community also calls for reinforcement of public health campaigns for the prevention and control of TSTC in the communities. About 40% of respondents in the community reported that they did not know about tapeworm infection in humans. These need to be enlightened about *T. solium* taeniosis through health education programs, as pork consumption was also high within the area and most of the slaughtered pigs were not inspected by veterinary officials. The lack of knowledge regarding transmission of TSTC in man and pigs could contribute to perpetuation of the tapeworm life cycle as knowledge about prevention was equally low. Furthermore, most of the respondents interviewed only attended primary education with an additional 15% not having attended any formal education. Thus, information on TSTC obtained through literature and formal education would be very low. A low level of education among rural communities has been known to increase the risk of *T. solium* infections and prevalence of NCC (26).

Regarding PCC, the majority of respondents reported having seen cysts (masese) in pork. Despite most of them (70%, 411) reporting knowing what cysts are, they could only describe how cysts looked like in pork meat mostly as “white watery things” or “small blisters in pork meat.” Most participants were also aware of the locations of *T. solium* cysticerci in the infected pigs with under the tongue and in muscles being the most commonly mentioned sites. When pig traders sell their pigs the commonest practice is to check for *cysticerci* under the tongue. Many also reported muscles as the other predilection site for cysticerci because they had seen infected pork meat before. Only a quarter of the respondents knew how pigs acquire the cysticerci. There is a strong misconception that feeding pigs with maize bran (gaga) left over from beer brew caused cysticerci in pigs. This is because of the similarity in appearance between the cysticerci (mases) and the peel from maize bran. This underscores the need for more health education to improve knowledge among communities. This lack of knowledge concerning transmission of PCC is not unique to our study but was also observed in Tanzania and Burkina Faso where only 33.7% and 6.2%, respectively, of the population knew that PCC was linked to pigs consuming human faeces (27,28). Overall, respondents were better informed about PCC than HCC. This is a common finding in most lower-income and middle-income countries where people are better informed about animal health than about their own health (21,27,29,30).

Regarding HCC more than half (57%, 334) of the respondents had heard about it in our study area but only 1% and 6% of these were knowledgeable about its transmission and prevention respectively. Similarly, while a good number of the participants (64%) knew the location of lesions for HCC, very few knew the signs and symptoms of HCC and NCC. Similar studies in Tanzania and Uganda found low knowledge regarding the signs and symptoms of HCC and NCC as well as its transmission (21,26,29). In the case of Tanzania most respondents had never even heard of HCC (21,31). Low awareness of HCC and NCC were also reported in the highly endemic areas of India and Mexico (32,33). In our case the community sensitization meetings played a role and contributed to the 39% of the population being knowledgeable about the signs and symptoms of NCC. Lack of knowledge of HCC and NCC could hinder efforts to control the most preventable cause of epilepsy in the sub-Saharan African region (6,7,34) as community awareness about a disease is important for its control (35).

Regarding epilepsy, the majority (more than 90%) of the participants had heard about it and more than half knew someone that had epilepsy. However, not many people knew that NCC is one of the causes of epilepsy. Only 8% of the participants responded that they knew the cause of epilepsy, but 17% of these cited witchcraft as the cause while 30% said it was caused by eating infected pork meat. Witchcraft was also cited as a cause of epilepsy in Tanzania by an even larger proportion (56%) of respondents (36). This misunderstanding with regards to the cause of epilepsy affects the choice of treatment as some would prefer to seek help from traditional healers if they suspected witchcraft as a cause. Fortunately, almost all participants reported that PWE should be taken to the hospital for treatment. A few, but significant proportion (7%) of the participants believed that epilepsy can be transmitted from one person to another through inhalation of flatus from a convulsing person or if one comes into contact with the sweat of a convulsing person. This belief that epilepsy can be transmitted from one person to another was also observed in a study conducted in a neighbouring district to our study area (17) as well as in a study conducted in Tanzania in which some respondents believed that epilepsy could be acquired if someone got into contact with saliva or froth from a person with epilepsy while having an epileptic seizure (36). These perceptions may lead to neglect of a person during a seizure as people will often fear to come into contact with a convulsing person. PWE are often stigmatized in many low-income and middle-income countries including Zambia, and the effects of this stigmatization can substantially decrease quality of life of PWE and their families (37,38). Educational interventions should thus emphasize that epilepsy is not transmissible from one person to another. Though epilepsy awareness and knowledge about treatment seems not too bad in our study area, the link between epilepsy and NCC has not been established by the local communities. In addition, there are persisting knowledge gaps to be addressed especially on the aetiology and transmission of epilepsy resulting in prejudice and stigma.

In this study, the majority of our participants obtained knowledge of TSTC in humans and pigs through community sensitisation programs conducted by *T. solium* researchers working in these communities some two years ago. However, despite those community sensitization meetings the level of knowledge about disease transmission and prevention in the majority was rather low.

Although community-based TSTC health education has been shown to have an impact on HCC and PCC (39,40), more robust behavioural efforts need to be implemented to improve community’s knowledge, attitudes and practices including the integration of the One Health approach. The fact that very few participants heard about human taeniosis and cysticercosis from the hospital or health facility could highlight a lack of knowledge among medical staff, as well as lack of appropriate diagnostic tools for TSTC. This lack of knowledge about TSTC infections in our study area is worrisome as it has been shown that lack of knowledge of populations about PCC and HCC leads to behaviours that facilitate the transmission and maintenance of *T. solium* infections (41,42).

Regarding attitudes and practices related to pig keeping and pork consumption, of the few participants that keep pigs, the majority raised their pigs through a free-range system. This is common practice within the region as it is said to be cheaper than confining the pigs in pig pens (16,17). Similar barriers to pig confinement have also been reported in other studies (39,43). Among those that reported consuming pork the majority preferred boiling their pork meat. However, 3.7% of the men preferred pork that was barbequed compared to that which was boiled. Also, 8.3% of the men said they consumed pork even if they saw cyst in it. This is common practice among men that drink alcohol. The dangers of eating inadequately roasted pork, particularly while drinking alcohol, was also identified in the Lao People’s Democratic Republic as a typically male risk factor requiring targeted intervention (44). There is thus a risk of *T. solium* eggs transmission from the men to the rest of the household and community as the main means of contamination remains the consumption of undercooked infected pork (43,45). Within the Eastern Province of Zambia men were found to be the most exposed to undercooked meat, translating in a higher taeniosis prevalence among them (46). With regards to sanitation and hygiene, while a good number of participants reported washing their hands always after using the toilet only 11% of the toilets had a hand washing facility (tippy tap) near to the toilet. Thus, education emphasizing the importance of maintaining hand washing stations with water and soap or ash, to prevent *T. solium* cysticercosis needs to be scaled up to reduce the risk of self-infection with *T. solium* eggs. Latrine usage, hand washing with soap and ensuring access to safe water represent behaviours that limit the burden of infectious diseases such as TSTC (47).

Overall, the KAP scores were very low within the Chiparamba area of Chipata district with the knowledge scores being slightly higher in comparison to attitude and practices scores. These findings reinforce the importance of increasing health education to strengthen KAP in Zambian communities. A further look at the knowledge scores revealed that males were significantly more associated with better knowledge about TSTC compared to females. This was also the case in terms of better practices observed among the males. Better knowledge among males compared to females was also observed in other studies conducted elsewhere. For example, in Uganda it was found that males had better knowledge about NCC compared to females (26). Similarly, in Tanzania, it was reported that male famers were significantly associated with better knowledge about PCC than their female counterparts (29).

The findings in this study are important for policy makers and all stakeholders that play a role in the control and elimination of TSTC. The poor knowledge, attitudes and practices regarding *T. solium* infections calls for an integrated and multisectoral approach in ensuring health education programs are developed together with affected communities and undertaken to improve awareness of this debilitating neglected tropical disease at different levels. The lack of meat inspection for slaughtered pigs and lack of knowledge regarding the importance of meat inspection requires that the community is engaged working together with the veterinary services and human health experts. Health education using a simple and meaningful health message for this complex zoonotic disease needs to be provided to many different stakeholders across disciplines and sectors as the life cycle of *T. solium* complicates the messaging (one worm causing three diseases) (48).

### Strengths and limitations

Our large sample size of over 500 households included in the study and the evaluation of comprehensive knowledge about TSTC are among the strengths to our study. The other strength is that the questions were administered in face-to-face interviews to each of the selected household members by enumerators conversant with the local language (Nsenga) common to both the Chewa and the Ngoni people of the area.

One limitation of our study is that we used a questionnaire with pre-specified answers. Thus, to correct for guessing of answers the enumerators were trained to ask the questions without stating the possible answers. The pre-specified answers also catered for the effect of using more than one enumerator during the study. Another limitation is that only one person per household was interviewed. This could have led to including respondents that were more or less knowledgeable about TSTC. The effect of this, however, could not be verified.

## Conclusion

This study revealed generally poor KAP among the community of Chiparamba in Chipata district of the Eastern Province of Zambia on TSTC. This poses a serious challenge to control and elimination of *T. solium* infections. Efforts to improve knowledge on TSTC transmission, signs and symptoms, prevention and management as well as its related attitudes and practices in Zambia need to be scaled up and this should be done in a One Health approach that includes the health of humans, animals and the environment they share in order to achieve control and elimination of *T. solium* infections. Considering that epilepsy is very prevalent in the whole of the Eastern Province and that the link of epilepsy to the different causes including NCC is poor, suggests that health workers may need more holistic training regarding TSTC, PCC, NCC and epilepsy.

## Data Availability

All relevant data are within the manuscript and its Supporting Information files.

## Acknowledgements

We thank the community of Chiparamba Rural Health centre catchment area who willingly offered their valuable time to participate in this study. We also wish to thank Phillip Chonya, Emmanuel Ngulube, Maxwel Masuku for their assistance with questionnaire survey data collection.

## Supporting information

S1 Table. Strobe checklist

S1 File. Supplementary tables.pdf

S2 File. Household questionnaire.doc

## Availability of data and material

All relevant data are within the manuscript and its supporting information files.

## Author Contributions

**Conceptualization:** Gideon Zulu, Kabemba E. Mwape, Tamara M. Welte, Isaac K. Phiri, Andrea S. Winkler.

**Data curation:** Gideon Zulu, Martin Simunza, Isaak K. Phiri.

**Formal analysis:** Gideon Zulu, Martin Simunza.

**Funding acquisition:** Chummy S. Sikasunge, Andrea S. Winkler.

**Investigation:** Gideon Zulu.

**Methodology:** Gideon Zulu, Kabemba E. Mwape, Tamara M. Welte, Wilborad Mutale, Isaac K. Phiri, Andrea S. Winkler.

**Project administration:** Gideon Zulu, Chummy S. Sikasunge.

**Resources:** Gideon Zulu.

**Software:** Gideon Zulu. Martin Simunza.

**Supervision:** Isaac K. Phiri, Andrea S. Winkler.

**Validation:** Gideon Zulu, Kabemba E. Mwape, Tamara M. Welte, Isaac K. Phiri, Andrea S. Winkler.

**Visualization:** Gideon Zulu, Isaac K. Phiri, Andrea S. Winkler.

**Writing – original draft:** Gideon Zulu.

**Writing – review & editing:** Kabemba E. Mwape, Tamara M. Welte, Martin Simunza, Alex Hachangu, Wilbroad Mutale, Mwelwa Chembensofu,, Chummy S. Sikasunge, Isaac K. Phiri, Andrea S. Winkler.

---

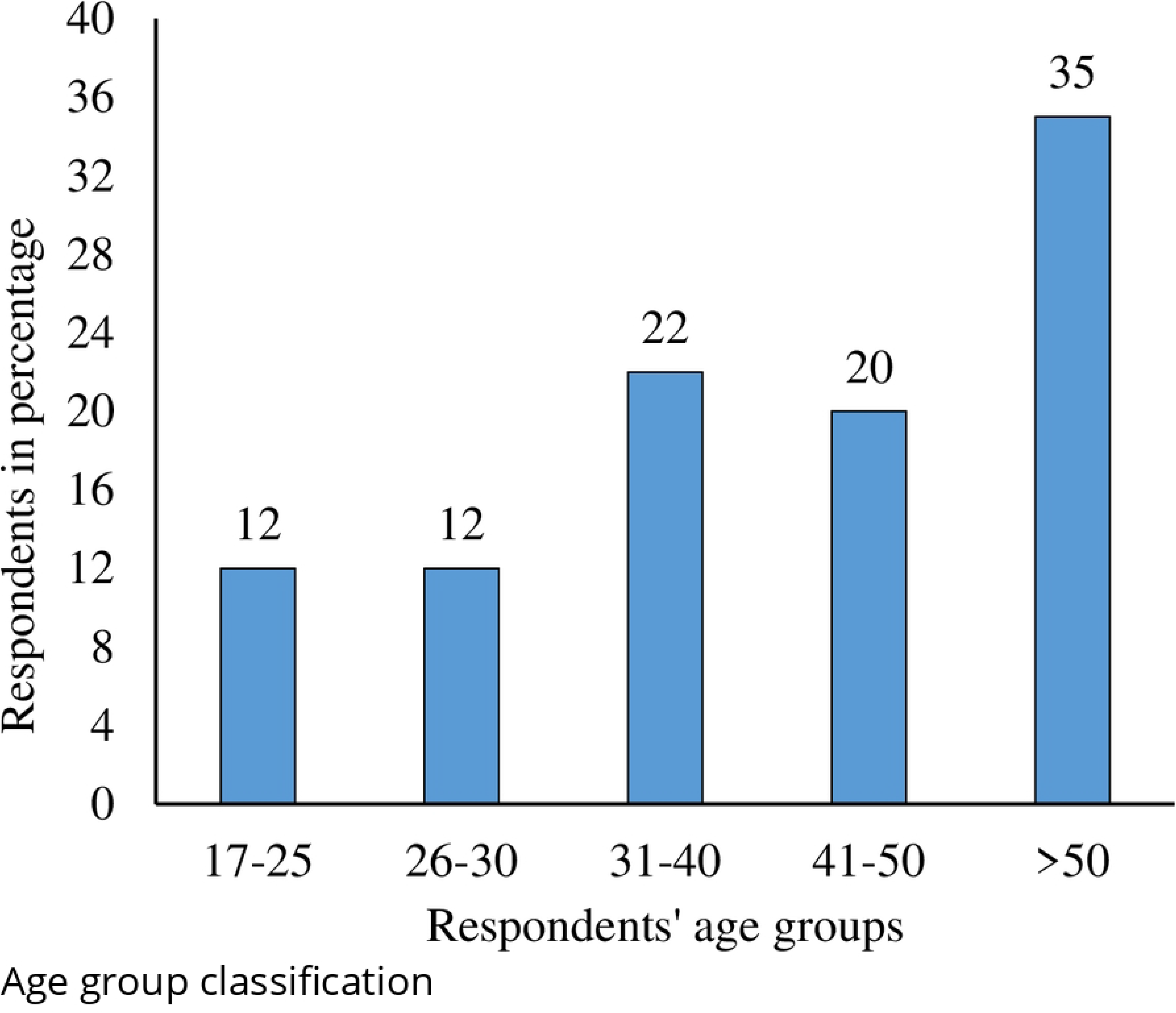

